# Indolent Presentations of Leukemic Lung Disease in Acute Myeloid Leukemia

**DOI:** 10.1101/2020.10.12.20211276

**Authors:** Yukiko Kunitomo, Seohyuk Lee, Chace C. Avery, Patricia L. Valda Toro, Avi J. Cohen, Solmaz Ehtashimi-Afshar, Peter A. Kahn, Alexa Siddon, Prajwal Boddu, Rupak Datta, Charles S. Dela Cruz, Samir Gautam

## Abstract

**Background:** Patients with active acute myelogenous leukemia (AML) are at risk for leukemic infiltration (LI) into the lung and acute tumor lysis pneumopathy (ATLP) following chemotherapy. Fulminant presentations of these leukemic lung diseases are well-described, but indolent forms have not yet been studied. Therefore, we sought to elucidate the clinical features of mild-to-moderate LI and ATLP.

**Methods:** A retrospective cohort analysis was performed on 51 hospitalized patients with AML, circulating blast count ≥3%, non-critical illness, and receipt of bronchoscopy between 2015-2019. Diagnoses of LI and ATLP were made via retrospective chart review by a multidisciplinary team of physicians.

**Results:** 19 cases of leukemic lung disease were identified: 14 with LI and 5 with ATLP. The clinical presentations closely resembled pneumonia, with the majority demonstrating respiratory symptoms (63%), hypoxemia (63%), fever (84%), and pulmonary opacities (100%). All patients were presumptively diagnosed with infection, leading to an average of 18 days of broad-spectrum antibiotic therapy and multiple instances of delayed chemotherapy in treatment candidates. Although most patients were near the end-of-life (90% died within 1 year), transitions to comfort care were infrequent (25%) and hospitalizations were protracted (median 25 days).

**Conclusions:** LI and ATLP are common yet under-recognized pulmonary complications in patients with active AML. When presenting indolently, these conditions are difficult to distinguish from lung infection, leading to missed diagnosis, inappropriate antibiosis, chemotherapy deferrals, and prolonged hospitalizations. Greater awareness and consensus definitions of LI and ATLP are therefore needed to improve care of this population.

## Introduction

Pulmonary complications are common in patients with acute myeloid leukemia (AML) and frequently life-threatening.^1,2^ Indeed, they occur in up to 80% of patients with AML and carry a 70% risk of intensive care unit (ICU) admission, with nearly 50% mortality.^1,3–8^ Due to the myelophthisic biology of AML and myeloablative chemotherapy used for induction, patients are often immunosuppressed and therefore susceptible to infection. Consequently, management of respiratory illness in this population focuses heavily on the identification and treatment of pneumonia.

Certain studies support the view that infection is the leading respiratory complication in patients with AML, but most are remote.^9–11^ More recent work indicates a growing proportion of non-infectious etiologies,^5^ including drug toxicities, diffuse alveolar hemorrhage (DAH), cardiogenic pulmonary edema, and leukemic lung disease.^12^ The reasons for this shifting epidemiology are unclear, but may relate to increased utilization of prophylactic antibiotics in leukemic patients and decreased incidence of bacterial pneumonia in the general population due to pneumococcal vaccination.^13–16^

Leukemic lung disease represents an important noninfectious cause of respiratory illness in patients with active AML. However, it is considered the rarest etiology in this population and therefore may be overlooked.^17^ Three manifestations of leukemic lung disease have been defined in the literature: (i) leukostasis, an emergent medical condition characterized by intravascular accumulation of blasts, increased blood viscosity, vascular inflammation, and multi-organ dysfunction;^18–20^ (ii) leukemic infiltration (LI) of blasts into the lung parenchyma; and (iii) acute tumor lysis pneumopathy (ATLP), an inflammatory lung injury precipitated by chemotherapy-induced lysis of blasts within the lung.^1,8,21,22^ Leukostasis is well-described, with clearly defined diagnostic criteria that facilitate prompt recognition and treatment.^18–20^ However, the clinical features of LI and ATLP in patients with AML have not been fully characterized.

The most extensive cohort of patients with leukemic lung disease to date was reported by Azoulay et al.^23^ These 20 cases, which were diagnosed based on clinical criteria, illustrate several important features including: (i) the proclivity for monocytic AML to invade the lung, (ii) the inconsistent association between circulating blast number and risk of LI, and (iii) the danger of ATLP after induction chemotherapy. This study also revealed the nonutility of bronchoalveolar lavage (BAL) in diagnosing LI, as blasts were usually absent from BAL fluid – a finding which corresponds with autopsies showing blasts in the interstitium and not the alveolar space.^24^ However, the study by Azoulay and colleagues was limited to patients in the ICU with rapidly progressive clinical courses. Subsequent case reports of LI and ATLP have focused on severe presentations as well.^25–28^

Therefore, we sought to characterize the clinical and radiographic features of indolent forms of LI and ATLP. To do so, we selected patients with AML admitted to non-ICU settings. We also required bronchoscopic evaluation to enable rigorous exclusion of lung infection, which represents the primary differential diagnosis in such patients. In the resulting cohort, we found LI and ATLP to be quite prevalent, but rarely recognized during hospitalization due to their clinical resemblance to lung infection.

## Methods

### Clinical Study Design

We performed a retrospective cohort study of adult patients ≥ 18 years of age who were admitted between 2015-2019 to Yale-New Haven Hospital (YNHH), a 1,541-bed tertiary care center in Connecticut, USA. To identify patients with active AML and mild-to-moderate respiratory illness, the following were required: (i) pathologic demonstration of AML on bone marrow biopsy; (ii) peripheral blast count ≥3% indicating active leukemia; (iii) receipt of bronchoscopy with BAL to evaluate respiratory illness; and (iv) initial admission to a non-ICU floor.

### Determination of respiratory illness etiology

We diagnosed patients with pulmonary infection if chart review identified any of the following: improvement of clinical status with empiric antimicrobial or antifungal therapy (as indicated by fever, hypoxemia, inflammatory biomarkers); positive serum fungal biomarkers; detection of respiratory viruses; or identification of bacteria or fungi by blood cultures, urine antigen testing, serum antigen testing (galactomannan or β-D-glucan), or lower respiratory cultures (except when deemed an obvious contaminant or colonizer by infectious disease and pulmonary consultants during hospitalization). Viruses were identified using a panel developed at YNHH as described previously^29^. BAL studies included bacterial cultures, fungal cultures, and viral nucleic acid detection. Specific testing for actinomyces spp., nocardia spp., *Legionella pneumophila*, acid-fast bacilli, and cytomegalovirus testing was performed for all patients. Empiric antiviral therapy (beyond acyclovir prophylaxis) was not used in the absence of positive diagnostic testing.

Diagnoses of noninfectious etiologies were made via chart review, with particular attention given to bronchoscopy results, drug exposures, autopsy and biopsy reports (where available), specialist consult notes, and clinical response to interventions. Cardiogenic pulmonary edema was diagnosed if the patient had (i) echocardiographic findings consistent with heart failure; (ii) evidence of volume overload on imaging or physical exam; and (iii) clinical improvement with diuresis. DAH was diagnosed if the BAL demonstrated increasingly hemorrhagic appearance on sequential aliquots. Sterile pneumonitis (drug-induced or graft versus host disease (GVHD)) was diagnosed if the patient had (i) biopsy confirmation; (ii) multi-organ involvement of active graft-versus-host-disease; or (iii) clear exposure to a culprit medication. Sepsis-related acute respiratory distress syndrome (ARDS) was diagnosed if the patient demonstrated (i) microbiological evidence of systemic infection and (ii) multi-organ dysfunction.

The diagnosis of leukemic lung disease was based on exclusion of all etiologies above *and* supporting clinical evidence for either LI or ATLP. The following clinical features were considered supportive of LI: (i) a rising blast count in the 60 days preceding admission in patients with known leukemia; (ii) improvement in respiratory status following chemotherapy in those receiving treatment during the hospital admission; (iii) progressive decline and death due to end-stage leukemia; and (iv) non-response to all antimicrobial or diuretic therapy. Features supporting a diagnosis of ATLP included: (i) elevated peripheral blast count prior to induction chemotherapy; (ii) clinical and/or radiographic decompensation following chemotherapy accompanied by a drop in blast count within 2 weeks of treatment. Review of patient charts started from the time from AML diagnosis, included the hospitalization, and continued until time of death. Of note, none of the patients had chronic underlying pulmonary disease or hypoxemia. Also, due to the exclusion of patients with severe clinical presentations, no patients had leukostasis.

To identify the final cohort with unambiguous leukemic lung disease, we required two additional criteria: comprehensive negative microbiological evaluation (respiratory, BAL, urinary and blood cultures, including potential contaminants or colonizers); and diagnostic confirmation by independent chart review by specialists in hematology, infectious disease, and pulmonology.

### Analysis of radiographic findings

All chest CT scans performed during hospitalization and the accompanying radiologist interpretations were reviewed. Descriptions of specific radiographic features and suspected diagnoses reported by the radiologists were extracted and tabulated.

### Data collection and analysis

Clinical data was obtained by the Joint Data Analytics Team at YNHH and medical record review. Data included demographics, comorbidities, vital signs, laboratory and imaging data, medication exposure, clinical interventions, and disposition. Data analysis was performed using Prism 8 (GraphPad Software, San Diego, USA) and the R software environment (version 3.6.0, Vienna, Austria). This study was approved by Yale’s institutional review board (#2000022465). Informed consent was waived due to the non-interventional study design.

## Results

### Identification of patients with indolent leukemic lung disease

We identified 71 patients with active leukemia admitted with non-critical illness who underwent bronchoscopy; 51 patients with pathologically confirmed AML were selected for further analysis (Figure E1). We excluded 24 patients based on microbiologic evidence of respiratory or systemic infection, including 3 patients suspected of having sepsis related ARDS. Another 8 patients were excluded for non-infectious respiratory diseases, resulting in a final cohort of 19 patients.

First, we sought to elucidate the etiologies of respiratory illness in the parent cohort of 51 AML patients (Table 1). These were determined by detailed post hoc chart review as described in the methods. Unexpectedly, we found that leukemic lung disease was the most common etiology, observed in 37% of the cohort (19 patients), while only 20% had definitive clinical and microbiological evidence of respiratory infection. Additional etiologies – including sterile pneumonitis (8%), sepsis-related ARDS (6%), DAH (4%), and cardiogenic pulmonary edema (4%) – were comparatively rare. The remaining 20% were found to have a high likelihood of leukemic lung disease, but were excluded due to the stringency of the diagnostic criteria (detailed in the methods).

**Table 1.**
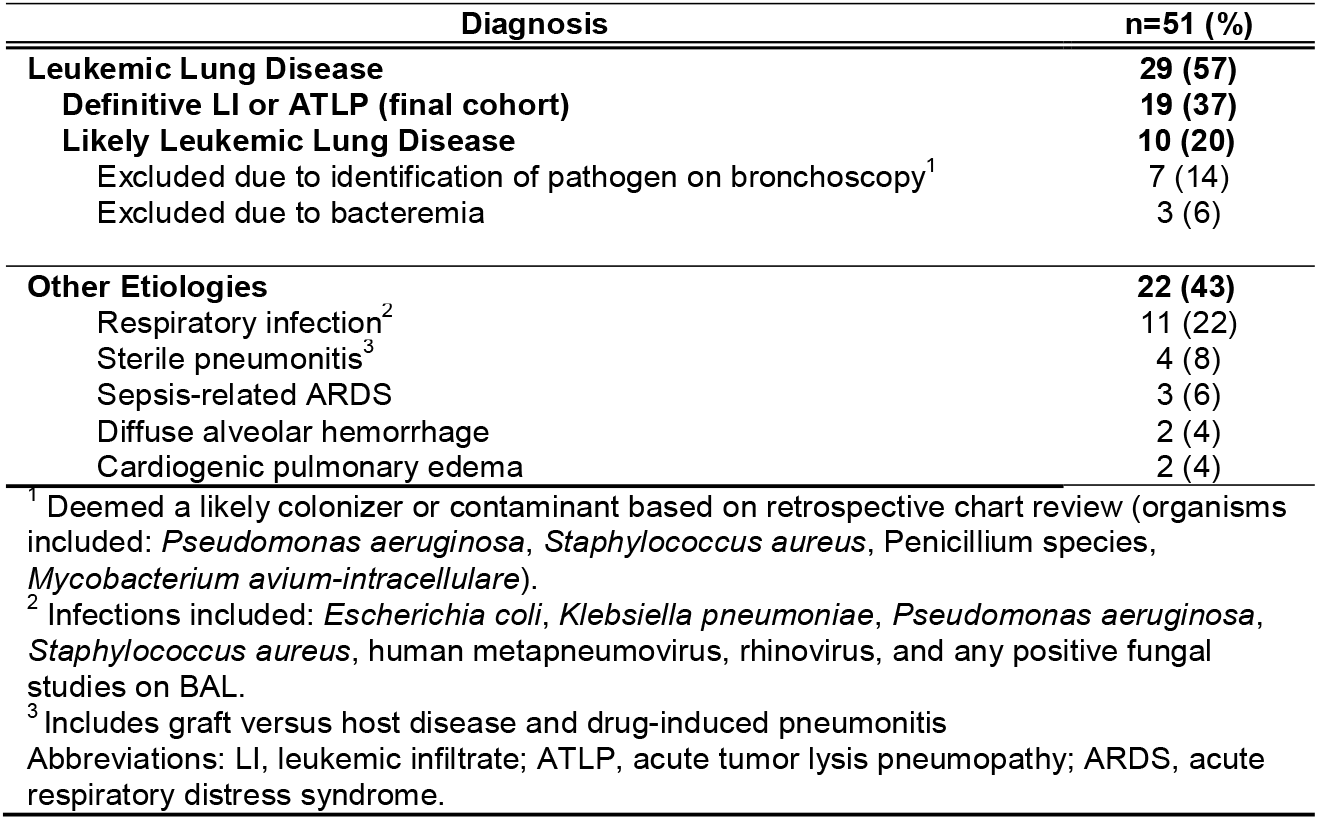
Etiologies of respiratory illness in AML patients who underwent bronchoscopy.

| Diagnosis | n=51 (%) |
| --- | --- |
| **Leukemic Lung Disease** | **29 (57)** |
| Definitive LI or ATLP (final cohort) | 19 (37) |
| Likely Leukemic Lung Disease | 10 (20) |
| Excluded due to identification of pathogen on bronchoscopy<sup>1</sup> | 7 (14) |
| Excluded due to bacteremia | 3 (6) |
| **Other Etiologies** | **22 (43)** |
| Respiratory infection<sup>2</sup> | 11 (22) |
| Sterile pneumonitis<sup>3</sup> | 4 (8) |
| Sepsis-related ARDS | 3 (6) |
| Diffuse alveolar hemorrhage | 2 (4) |
| Cardiogenic pulmonary edema | 2 (4) |

<sup>1</sup> Deemed a likely colonizer or contaminant based on retrospective chart review (organisms included: *Pseudomonas aeruginosa*, *Staphylococcus aureus*, *Penicillium* species, *Mycobacterium avium-intracellulare*).
<sup>2</sup> Infections included: *Escherichia coli*, *Klebsiella pneumoniae*, *Pseudomonas aeruginosa*, *Staphylococcus aureus*, human metapneumovirus, rhinovirus, and any positive fungal studies on BAL.
<sup>3</sup> Includes graft versus host disease and drug-induced pneumonitis
Abbreviations: LI, leukemic infiltrate; ATLP, acute tumor lysis pneumopathy; ARDS, acute respiratory distress syndrome.

### Clinical presentation and hospital course

The final cohort of 19 patients was subdivided into new AML (N-AML) and relapsed or refractory AML (RR-AML), which was defined by any receipt of induction chemotherapy prior to hospitalization. The clinical presentations and courses of individual patients are diagrammed in Figure E2, while summative data of the cohort is presented in Tables 2 and 3. Per study design, disease severity and blast counts were relatively low, as the average WBC count was 22×10^3^/μL and no patients exceeded 100×10^3^/μL. Based on current World Health Organization guidelines,^30^ all patients had non-acute promyelocytic leukemia (Table 1), with the most common subtype being AML with myelodysplasia related changes (47%).

**Table 2.**
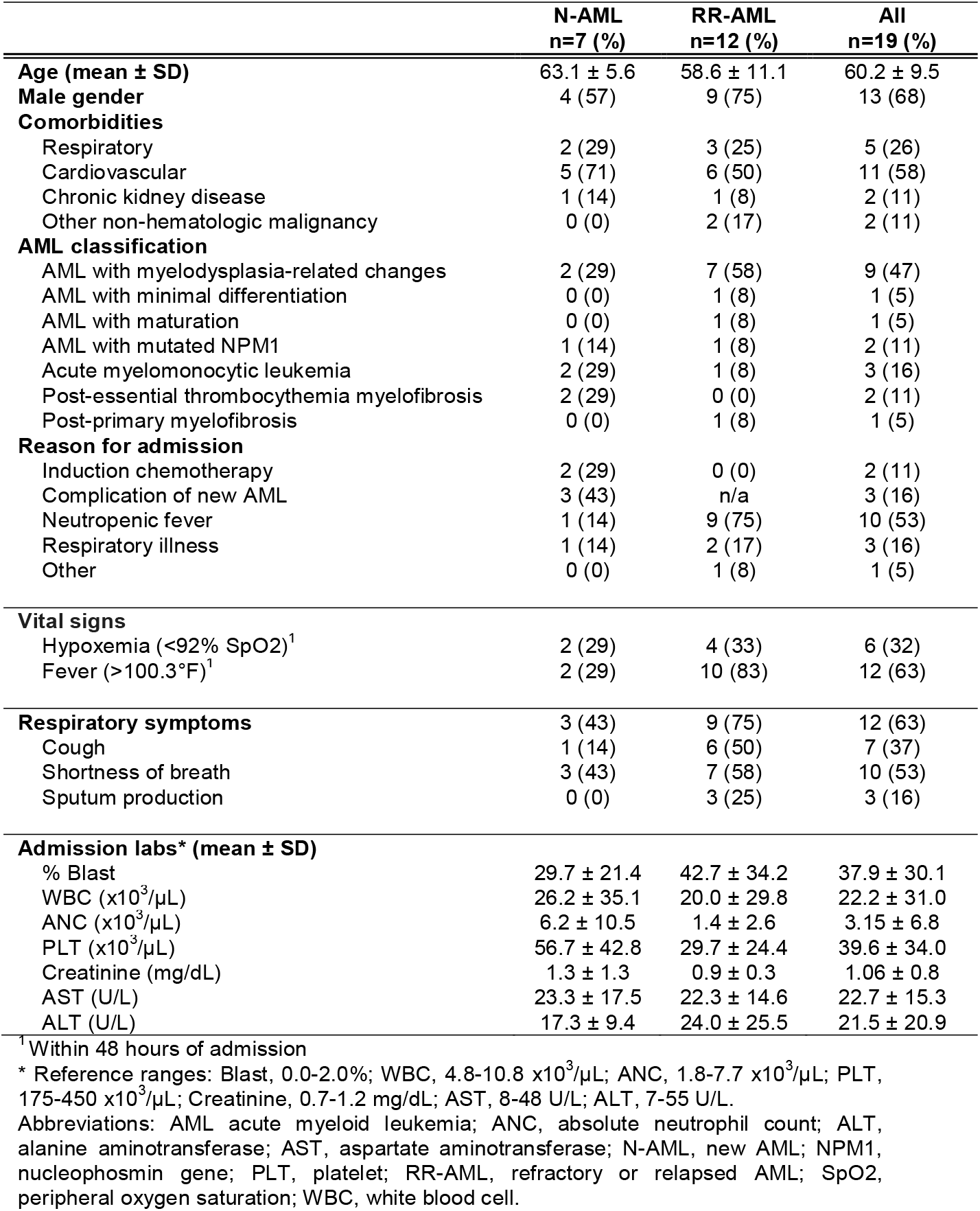
Patient demographics and notable clinical and laboratory findings on admission.

**Table 3.**
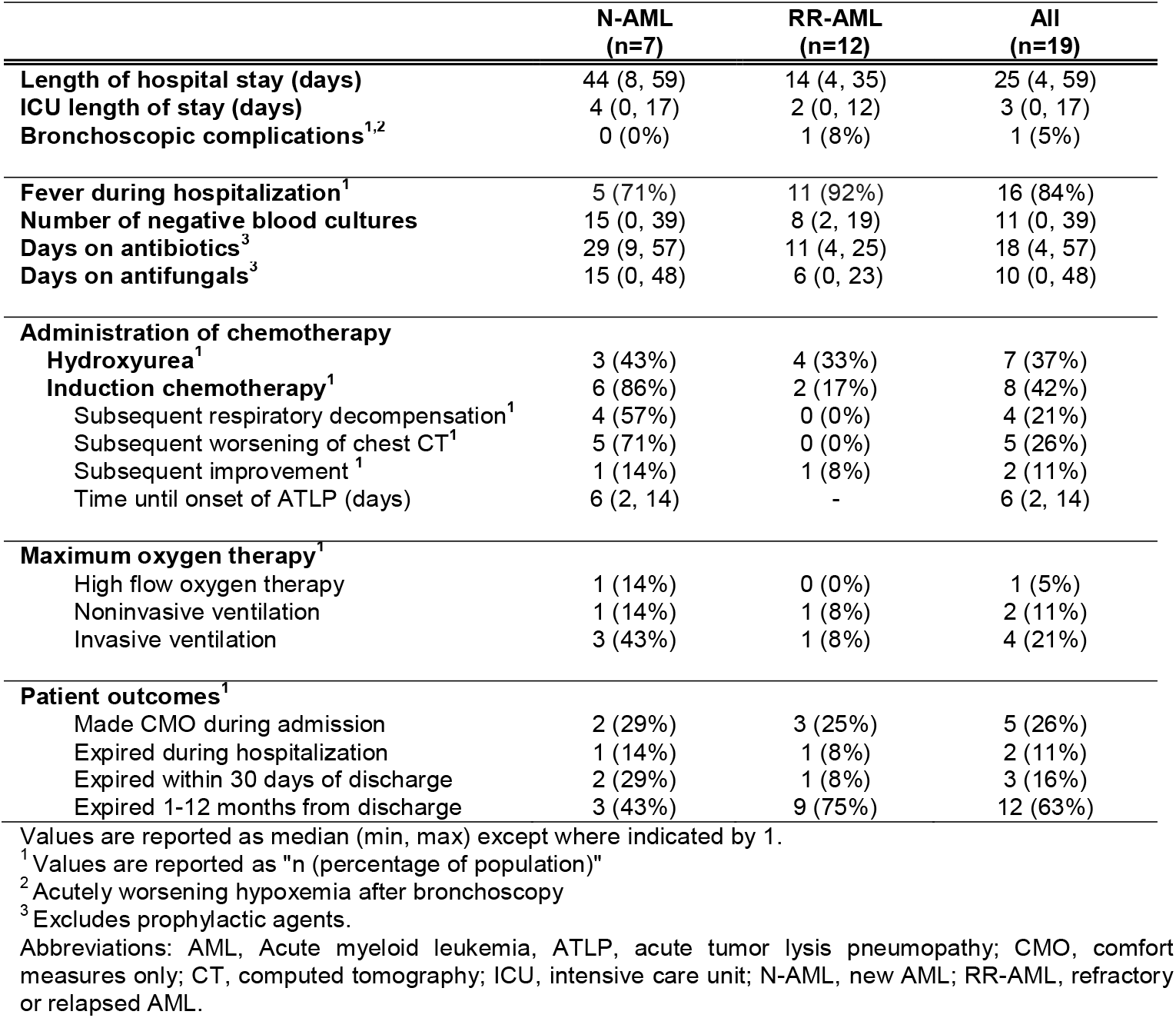
Major clinical events and interventions during hospitalization.

#### N-AML

Of the 7 patients with N-AML, 6 were treated with chemotherapy, while 1 had treatment deferred due to concern for infection. One of the six receiving chemotherapy demonstrated marked improvement in respiratory status – a phenomenon also observed previously in three patients with LI by Kovalski et al.^31^ The remaining 5 experienced worsening hypoxemia and/or radiographic infiltrates due to ATLP, although respiratory decompensation was attributed to infection in all cases at the time of hospitalization.

In contrast to the patients reported by Azoulay et al, all of whom developed life-threatening ATLP within 48h of chemotherapy,^23^ we noted a marked delay in onset. Indeed, the average time to development of ATLP was 6 days, with one case manifesting 14 days after therapy. This difference may relate to the lower peripheral blast counts in our cohort. However, despite the modest blast counts and milder initial clinical presentations, the majority of ATLP cases still developed severe respiratory decompensation requiring ICU transfer (4/5 patients).

Interestingly, we also noted a subset of patients who showed negligible pulmonary signs and symptoms upon admission but developed severe ATLP after chemotherapy (2/5 patients). We suspect that these patients had ‘subclinical’ leukemic infiltration on admission, though this went unrecognized because of a lack of symptoms and imaging prior to chemotherapy.

#### RR-AML

Patients with RR-AML and leukemic lung disease demonstrated a highly characteristic rise in peripheral blast count during the weeks-to-months prior to hospitalization (Figure E2). They usually presented with fever (83%), respiratory symptoms (75%), and radiographic infiltrates (100%) that were caused by LI but attributed to infection at the time of hospitalization. In at least one case, this led to deferment of chemotherapy. Also, as in the N-AML group, we observed one instance of marked clinical improvement after chemotherapy. Finally, we noted that only 3/12 patients were transitioned to comfort or palliative care despite their end-stage disease (11/12 died within one year), largely based on the perception that their decompensation was due to infection rather than progressive leukemia.

#### General features

All 19 patients with leukemic lung disease received heavy exposure to antimicrobial agents (Table 2), with a mean duration of 18 days of broad-spectrum antibiotics (maximum of 57 days) and 10 days of treatment-dose antifungals (maximum of 48 days). The average number of blood cultures during admission was 11 with a maximum of 39 (all negative per inclusion criteria). Length of stay was quite prolonged, with a mean of 25 and maximum of 35 days.

### Radiographic findings

Two prior case series, which recruited 1 and 6 patients with AML, respectively, have reported the radiographic features of LI.^32,33^ The larger study suggested that septal and/or bronchial wall thickening may be a characteristic finding.^33^ However, in our cohort of patients with indolent LI, this was only observed in 25% (Table 4). Indeed, we found the radiographic appearance of LI to be highly variable and lacking in features that distinguished it from infection and other etiologies (Figure 2A-D). In contrast, ATLP predominantly manifested with ground glass opacities (80%), as exemplified by one patient who underwent CT scans before and after chemotherapy (Figure 2E-F). In both LI and ATLP, bilateral lung involvement was common. Also, in both diseases, radiologists frequently indicated concern for pneumonia, with more than half explicitly identifying infection as the leading diagnostic suspicion.

**Table 4.**
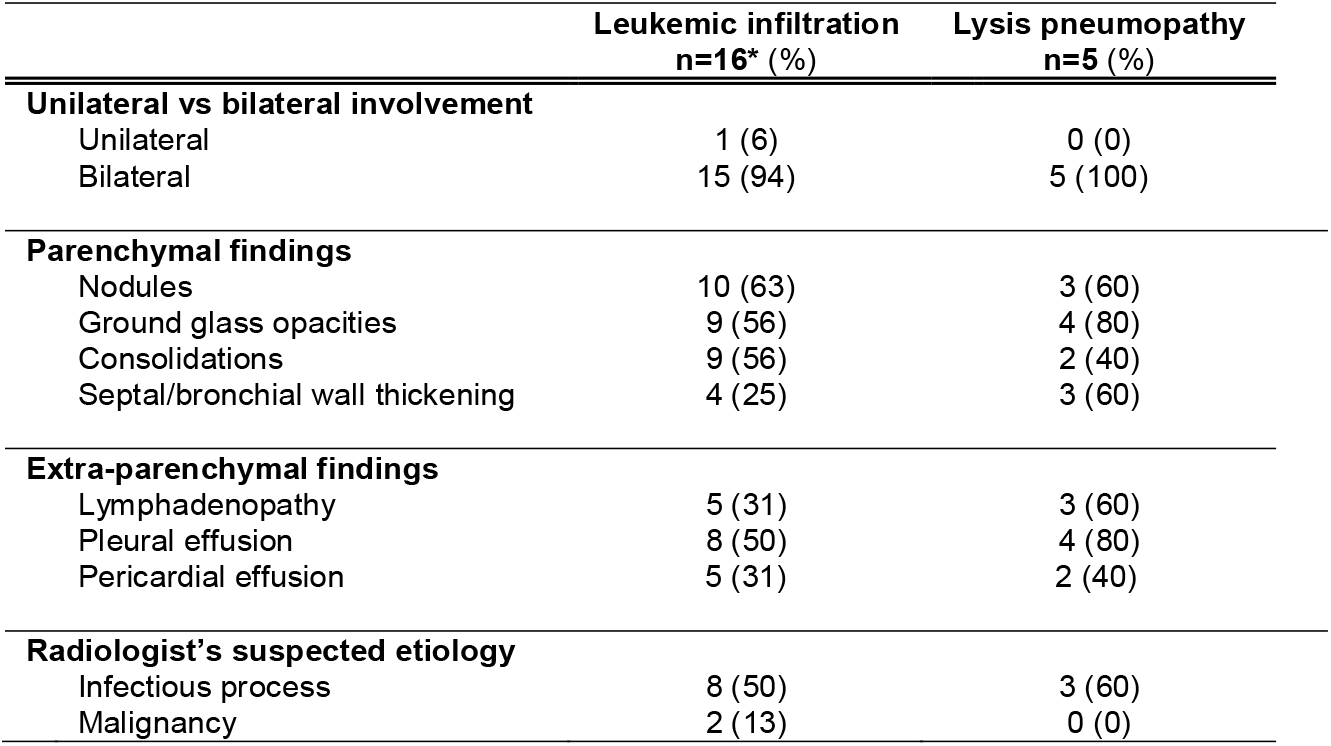
Summary of major CT chest findings.

|  | **Leukemic infiltration**<br>**n=16\* (%)** | **Lysis pneumopathy**<br>**n=5 (%)** |
| --- | --- | --- |
| **Unilateral vs bilateral involvement** |  |  |
| Unilateral | 1 (6) | 0 (0) |
| Bilateral | 15 (94) | 5 (100) |
| **Parenchymal findings** |  |  |
| Nodules | 10 (63) | 3 (60) |
| Ground glass opacities | 9 (56) | 4 (80) |
| Consolidations | 9 (56) | 2 (40) |
| Septal/bronchial wall thickening | 4 (25) | 3 (60) |
| **Extra-parenchymal findings** |  |  |
| Lymphadenopathy | 5 (31) | 3 (60) |
| Pleural effusion | 8 (50) | 4 (80) |
| Pericardial effusion | 5 (31) | 2 (40) |
| **Radiologist's suspected etiology** |  |  |
| Infectious process | 8 (50) | 3 (60) |
| Malignancy | 2 (13) | 0 (0) |

## Discussion

The cohort presented here offers new insight into LI and ATLP, two common yet underappreciated pulmonary complications of active AML. Focusing on non-ICU patients, we provide the first descriptions of indolent forms of these diseases. Taken together with the work of Azoulay et al on severe LI and ATLP,^23^ our findings reveal a broad continuum of clinical presentations ranging from mild to fulminant. In fact, we also observed cases of LI in asymptomatic patients, which we designate subclinical LI. Thus, clinicians should be vigilant for the possible presence of LI in any patient with active AML, and for the risk of ATLP when treating such patients with chemotherapy – even if asymptomatic on presentation. Diagnosis of ATLP, in turn, should be strongly suspected in patients who develop respiratory symptoms after induction.

As stated above, severe leukemic lung diseases such as leukostasis are often identified by clinicians, in part based on the dramatic elevations in WBC that usually accompany these conditions (i.e. hyperleukocytosis). In our cohort with indolent disease, however, diagnoses of LI and ATLP were missed with striking regularity. Instead, respiratory illness was invariably attributed to infection, resulting in delays in chemotherapy for treatment candidates and palliative care transitions for noncandidates. This occurred despite comprehensive testing including bronchoscopy, as well as multidisciplinary consultation with specialists in pulmonology, infectious disease, chest radiology, and hematology.

The presumption of lung infection in these patients is partly explained by the limited specificity of diagnostic criteria for pneumonia – i.e. respiratory symptoms, systemic signs of infection (e.g. fever), and radiographic infiltrates – all of which occur commonly in leukemic lung disease.^34,35^ Conversely, establishing a definitive diagnosis of LI represents a challenge as well. This stems from the risks of obtaining biopsies for pathological confirmation,^36,37^ nonutility of BAL studies,^23^ inconsistent association with peripheral blast number,^23,38^ and non-specificity of radiographic findings.^31,33,38^ Accordingly, LI is a diagnosis of exclusion in most cases.

Since LI and pneumonia are virtually indistinguishable by standard clinical, radiographic, and pathological measures, a more tailored diagnostic approach is required. We would advocate for the following: (i) prompt radiographic and comprehensive microbiological evaluation including bronchoscopy; (ii) administration of empiric broad-spectrum antimicrobial therapy (as well as diuresis in appropriate patients); and (iii) diagnostic re-evaluation after finalization of infectious studies and completion of a defined course of empiric treatments. Negative microbiology and non-response to therapy may reasonably justify antimicrobial de-escalation and a tentative diagnosis of LI, especially in the presence of supportive features such as a concurrence between respiratory manifestations and rising blast count. In such cases, consideration should be given to cytoreduction for treatment candidates, especially given the marked clinical improvement observed after chemotherapy in multiple patients in our cohort, and previously by Kovalski et al.^31^

However, as demonstrated by 5 patients in this study (4 of whom required ICU transfer), the risk for precipitating severe ATLP is significant even with relatively low peripheral blast counts, although onset may be delayed. Therefore, routine chest CT imaging may be indicated prior to chemotherapy to evaluate for LI. In those diagnosed with LI, gradual cytoreduction (e.g. with hydroxyurea) may be preferable to immediate induction. Prophylactic steroids may further mitigate the risk of ATLP as indicated by the work of Azoulay et al,^39^ and close monitoring for at least 7 days after chemotherapy is warranted based on the delayed onset of ATLP in our cohort. Meanwhile, the development of LI in treatment-refractory AML may be viewed as evidence of end-stage disease and justification for transition to palliative care.

The prevalence of LI and ATLP in our cohort represents another notable finding of the present study. In contrast to the conventional view that leukemic lung disease represents the rarest cause of respiratory illness in AML, it was strongly suspected in the *majority* of our parent cohort (57%). These results are consistent with autopsy reports showing a 18-64% prevalence of LI in end-stage leukemics,^11,17,40^ and emphasize the need to maintain a high index of suspicion for this diagnosis in patients with active AML. Given the frequency of LI in this population, it may also present concurrently with infection, though our study does not address this question.

The pathophysiology of LI and ATLP remains poorly understood, but recent advances in the biology of myeloid cells such as neutrophils may offer useful insight. For instance, myeoloblasts and neutrophils both demonstrate a tendency to marginate in the pulmonary vasculature,^40–44^ and the chemokine receptor CXCR4 is now recognized to be important for this effect in neutrophils.^45^ Given the association between CXCR4 and extramedullary infiltration, as well as heightened expression on monocytic leukemia (which tends to involve the lung), it is plausible that CXCR4 also mediates retention of blasts in the pulmonary vasculature.^46,47^ Extravasation, in turn, may proceed by mechanisms described by Stucki et al: i.e. activation of endothelial cells by blast-derived cytokines, upregulation of adhesion molecules on the endothelium, and transmigration.^48^ Finally, induction of ATLP during cytolytic chemotherapy could result from necrotic release of danger-associated molecular patterns (DAMPs) from blasts within the pulmonary vasculature and/or parenchyma, leading to pulmonary inflammation (and ground glass opacities on CT). While further studies are needed to confirm these postulations, it is tempting to speculate that CXCR4 inhibition prior to chemotherapy could promote neutrophil decampment from the lung and thus decrease the risk ATLP.

The principal limitation to our study is the absence of pathologic confirmation of leukemic lung involvement. This represents a pervasive obstacle to diagnosing LI and derives from the risk of pulmonary hemorrhage when performing transbronchial biopsy in patients with AML, who are often severely thrombocytopenic. While autopsy represents a viable alternative for post hoc confirmation, it is usually declined by our patient population. Nevertheless, we believe the strict requirement for negative bronchoscopic studies, rigorous retrospective analysis of cases by independent clinicians from multiple specialties, and provision of objective patient clinical course diagrams (Figure E2) strongly support our diagnoses of leukemic lung disease. In fact, given the potential for LI to present concurrently with infection, it may be better diagnosed by clinical features rather than pathology alone, as the latter may underestimate the contribution of coinfection. Finally, prior studies which serve as the foundation for the literature in the field have demonstrated the validity of diagnosing LI by clinical criteria alone.^23^

Other limitations include the retrospective and single center aspects of the study, which may have introduced bias related to patient selection for bronchoscopy (one of the central inclusion criteria). Indeed, there may have been patients with clinically obvious infection who were not referred for bronchoscopy, therefore increasing the relative proportion of patients diagnosed with leukemic lung disease in our cohort. However, since we strongly emphasize bronchoscopy in the evaluation of all patients with active AML and respiratory illness at our institution, this likely had minimal effect on the present study. Finally, we restricted our analysis to AML, both to maintain homogeneity in our study cohort, and to address the perception that infection is particularly prevalent in this population. As such, further studies will be necessary to see how well the clinical features reported here apply to other forms of leukemia.

Further work is also necessary to elucidate the biological mechanisms of LI in order to develop improved diagnostics, treatments, and strategies to prevent ATLP. Such studies, along with heightened clinical awareness of leukemic lung disease, should help improve antibiotic stewardship in patients with AML, encourage chemotherapy administration where appropriate, and expedite care de-escalation in treatment noncandidates. Fortunately, efforts to standardize the management of respiratory illness and reduce antibiotic overuse in AML and other immunocompromising diseases are now underway.^49^

**Figure 1.**
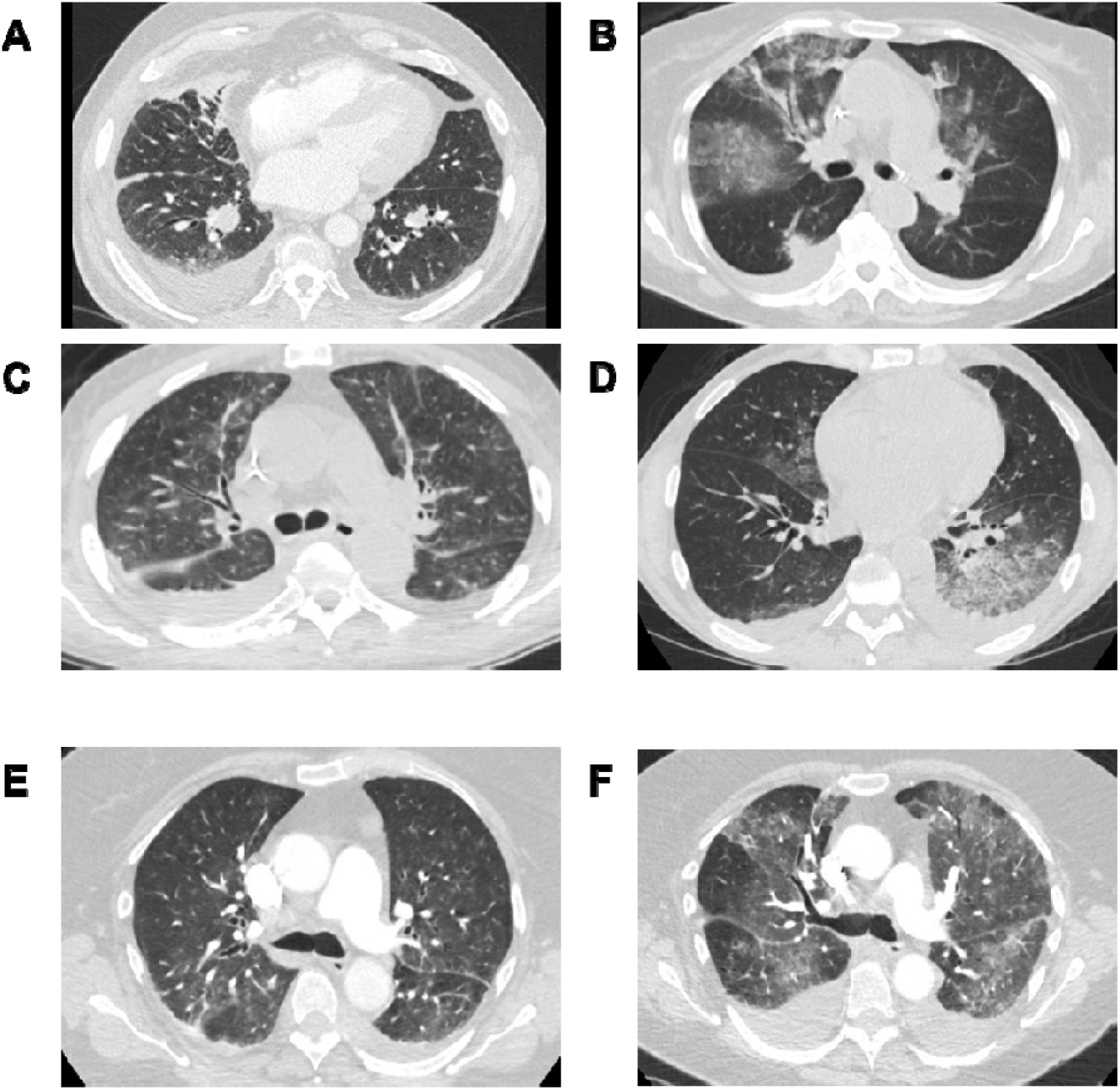
Representative CT chest scans. A, Nodules; B, ground glass opacities; C, septal/bronchial wall thickening; D, consolidation. E, Pre-chemotherapy admission CT; F, post chemotherapy CT.

## Data Availability

Deidentified data related to this study is available upon request addressed to the corresponding author

