## Supplemental Figure E2 for "Indolent Presentations of Leukemic Lung Disease in Acute Myeloid Leukemia"

**Figure E2: Patient course diagrams.** New AML (N-AML_1-6) patient courses cover duration of hospital admission. Relapsed AML (RR-AML_1-12) patient courses include blast data prior to admission (up to 6 months prior) and duration of hospital admission.

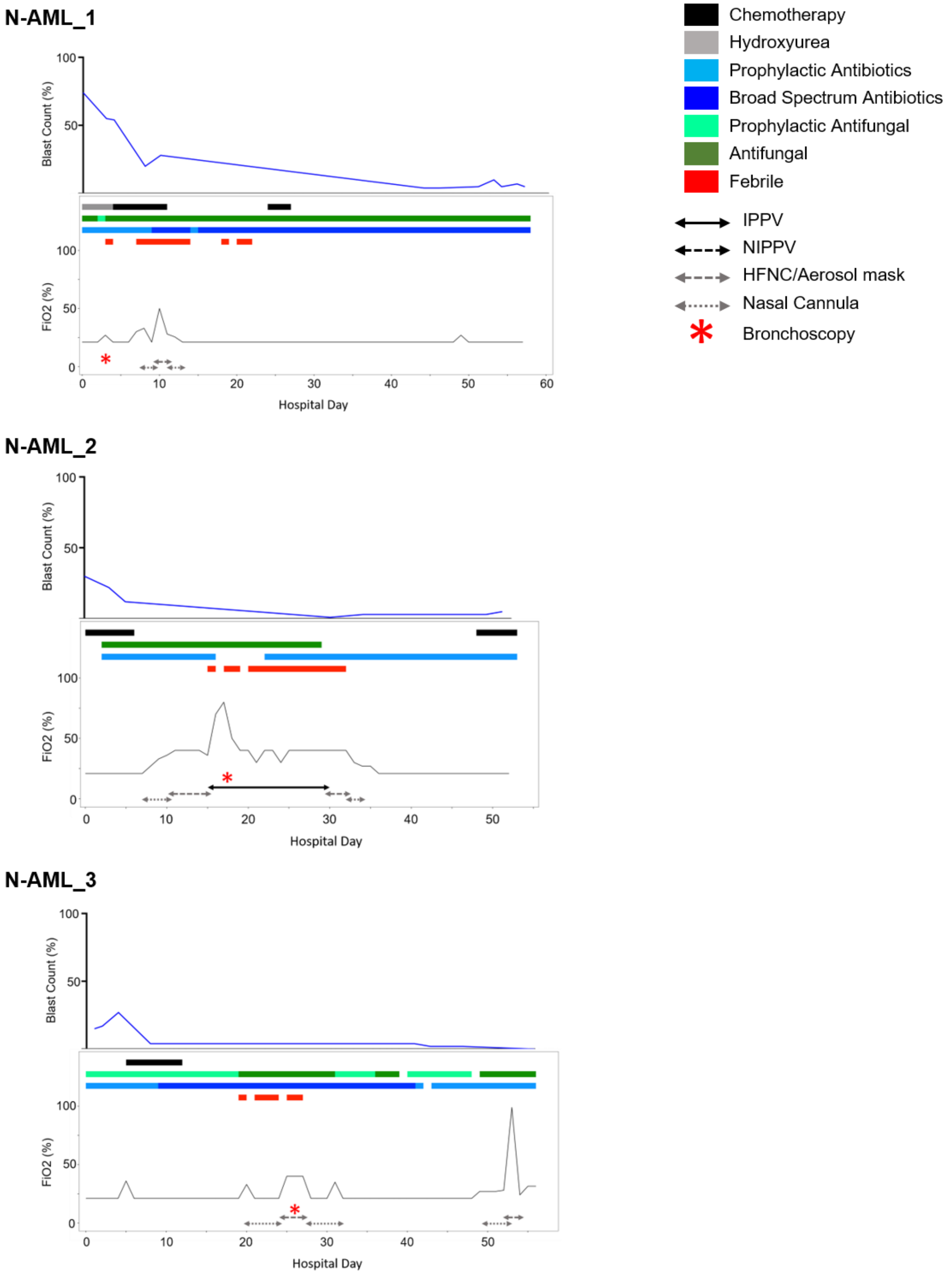

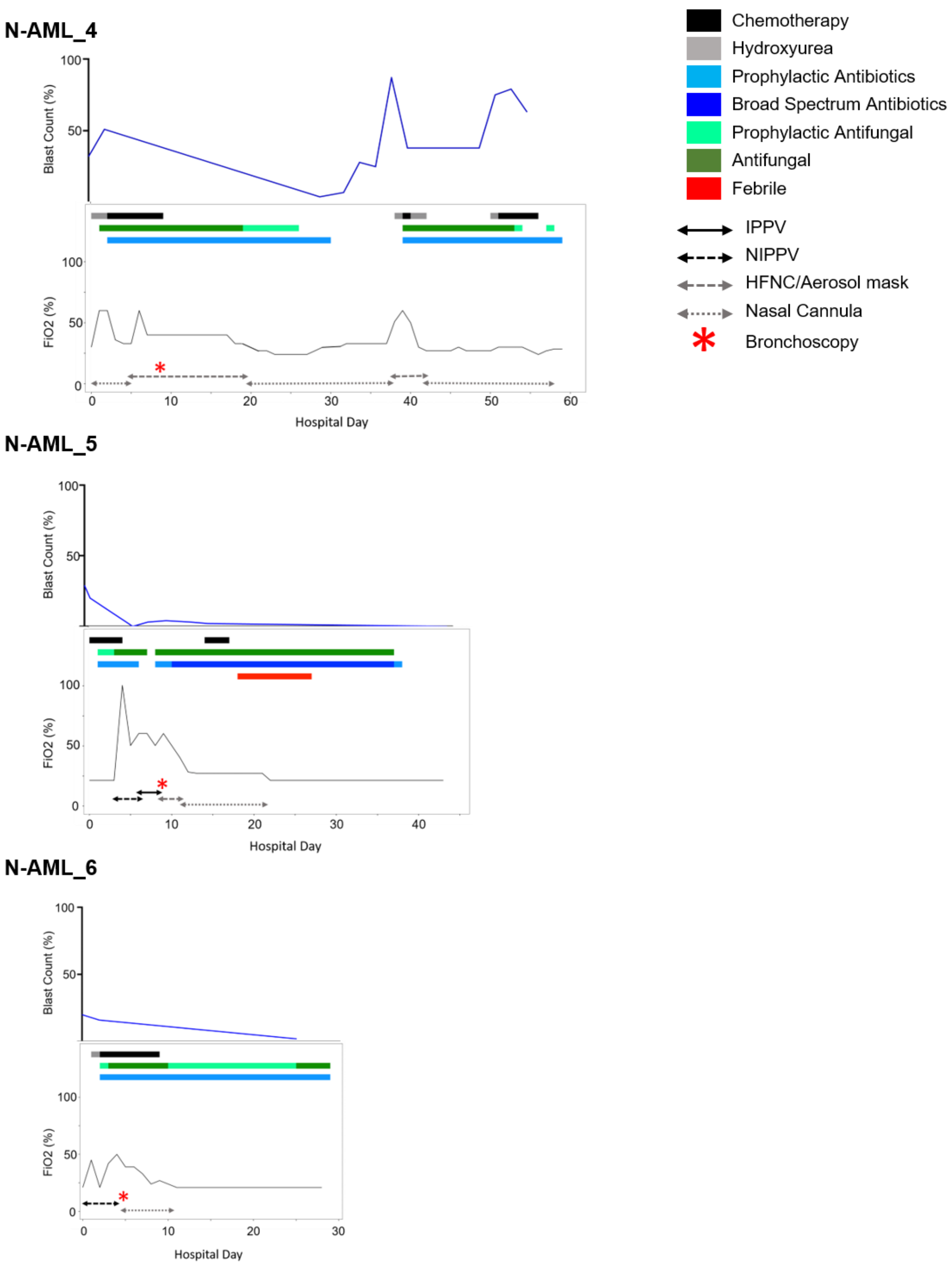

N-AML_4
Blast Count (%)
100
50
FiO2 (%)
100
50
0
0
10
20
30
40
50
60
Hospital Day
N-AML_5
Blast Count (%)
100
50
FiO2 (%)
100
50
0
0
10
20
30
40
Hospital Day
N-AML_6
Blast Count (%)
100
50
FiO2 (%)
100
50
0
0
10
20
30
Hospital Day
Chemotherapy
Hydroxyurea
Prophylactic Antibiotics
Broad Spectrum Antibiotics
Prophylactic Antifungal
Antifungal
Febrile
IPPV
NIPPV
HFNC/Aerosol mask
Nasal Cannula
Bronchoscopy

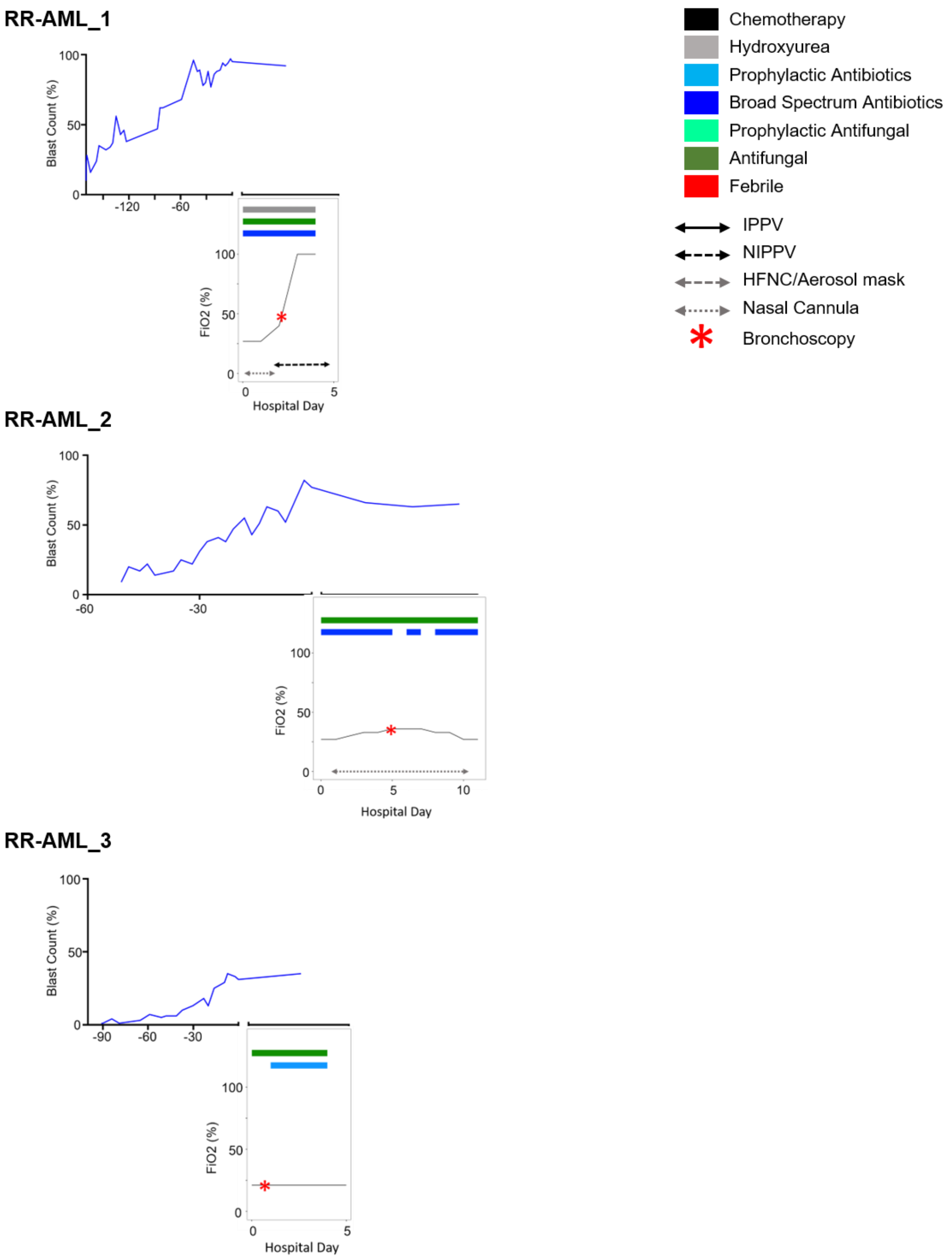
RR-AML_1
Blast Count (%)
100
50
0
-120
-60
FiO2 (%)
100
50
0
0
5
Hospital Day
RR-AML_2
Blast Count (%)
100
50
0
-60
-30
FiO2 (%)
100
50
0
0
5
10
Hospital Day
RR-AML_3
Blast Count (%)
100
50
0
-90
-60
-30
FiO2 (%)
100
50
0
0
5
Hospital Day
Chemotherapy
Hydroxyurea
Prophylactic Antibiotics
Broad Spectrum Antibiotics
Prophylactic Antifungal
Antifungal
Febrile
IPPV
NIPPV
HFNC/Aerosol mask
Nasal Cannula
Bronchoscopy

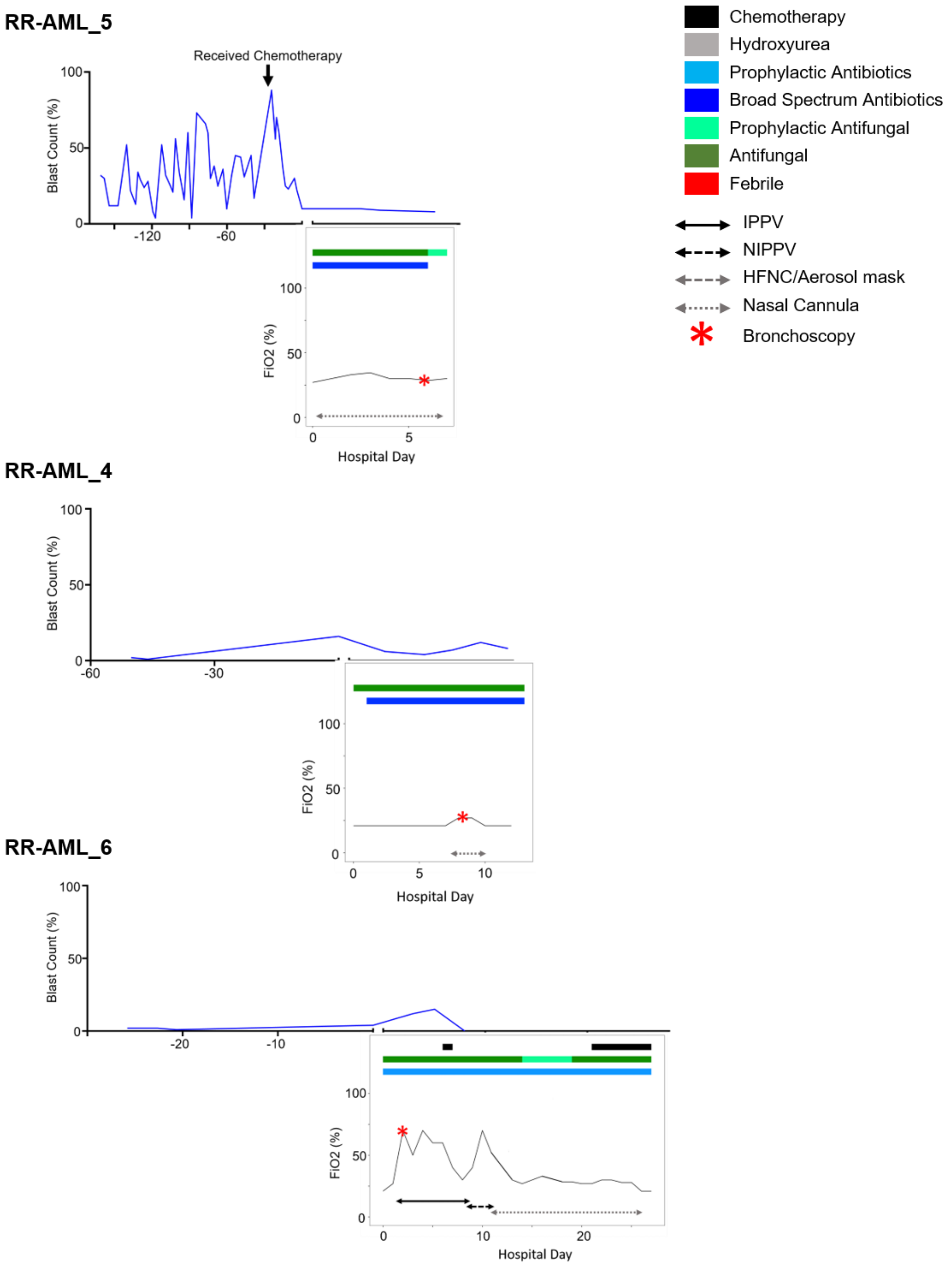
RR-AML_5
Received Chemotherapy
Blast Count (%)
100
50
0
-120
-60
FiO2 (%)
100
50
0
0
5
Hospital Day
RR-AML_4
Blast Count (%)
100
50
0
-60
-30
FiO2 (%)
100
50
0
0
5
10
Hospital Day
RR-AML_6
Blast Count (%)
100
50
0
-20
-10
FiO2 (%)
100
50
0
0
10
20
Hospital Day
Chemotherapy
Hydroxyurea
Prophylactic Antibiotics
Broad Spectrum Antibiotics
Prophylactic Antifungal
Antifungal
Febrile
IPPV
NIPPV
HFNC/Aerosol mask
Nasal Cannula
Bronchoscopy

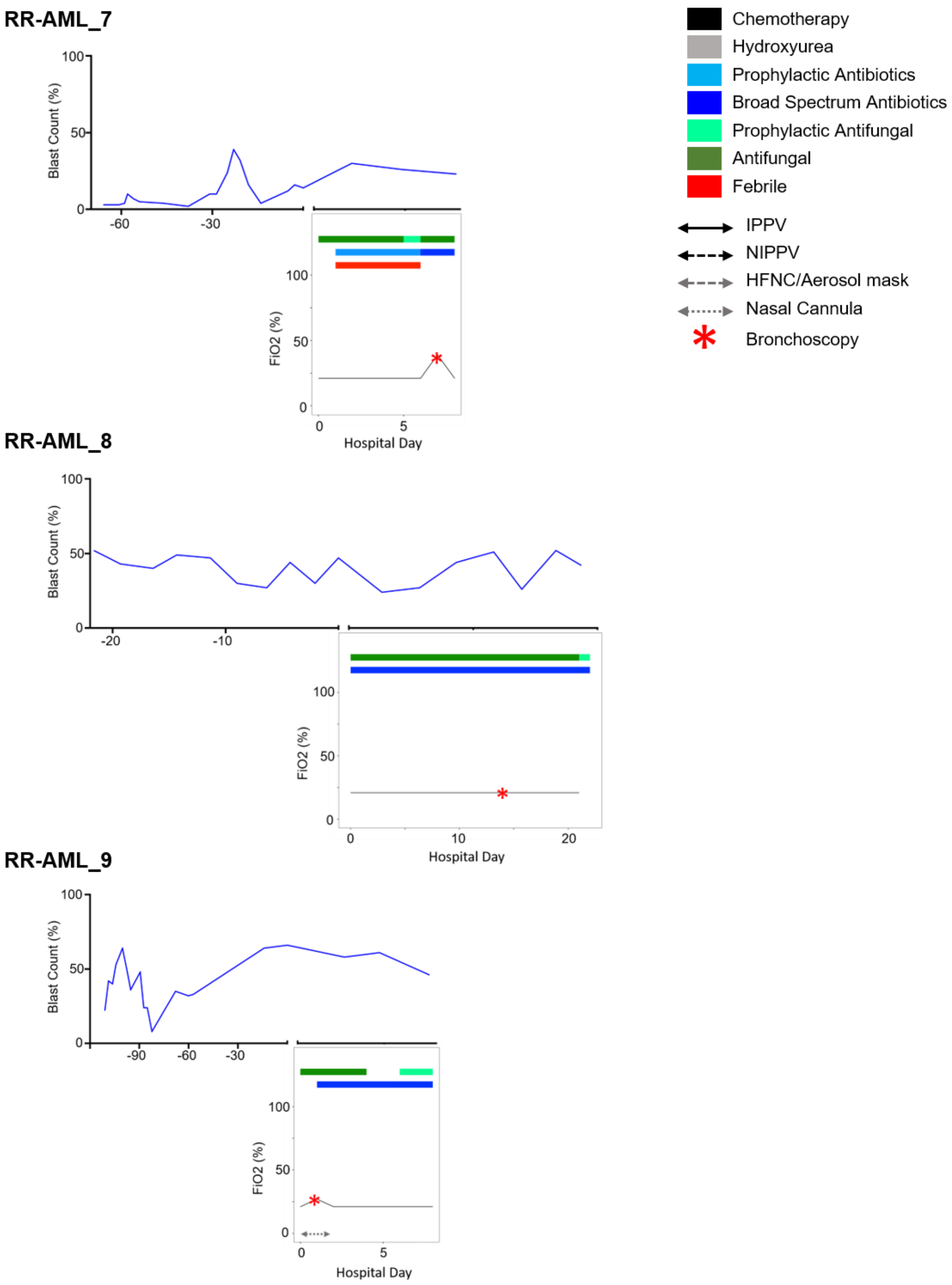

RR-AML_7
Blast Count (%)
100
50
0
-60
-30
FiO2 (%)
100
50
0
0
5
Hospital Day
RR-AML_8
Blast Count (%)
100
50
0
-20
-10
FiO2 (%)
100
50
0
0
10
20
Hospital Day
RR-AML_9
Blast Count (%)
100
50
0
-90
-60
-30
FiO2 (%)
100
50
0
0
5
Hospital Day
Chemotherapy
Hydroxyurea
Prophylactic Antibiotics
Broad Spectrum Antibiotics
Prophylactic Antifungal
Antifungal
Febrile
IPPV
NIPPV
HFNC/Aerosol mask
Nasal Cannula
Bronchoscopy

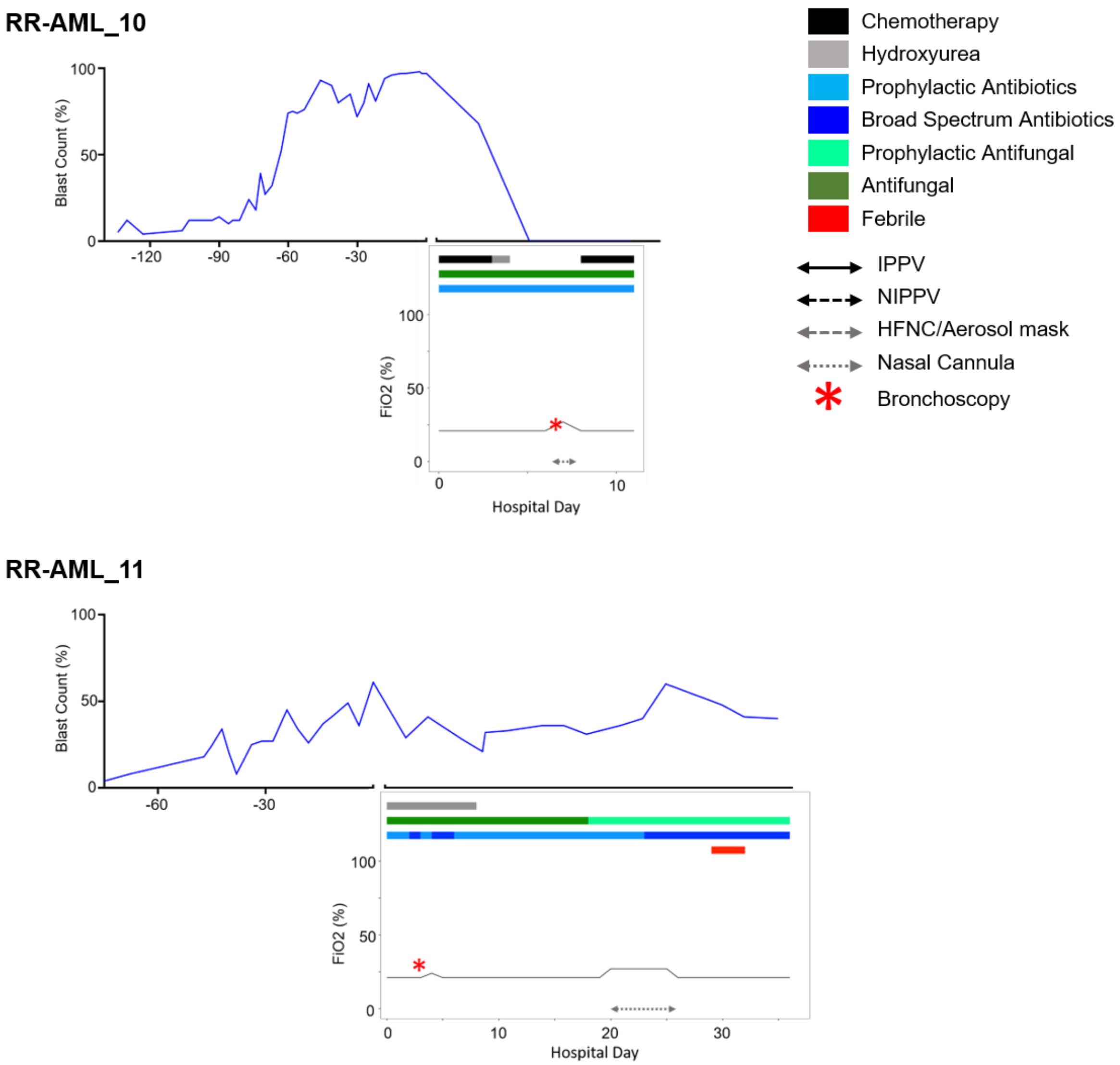

RR-AML_10
Blast Count (%)
100
50
0
-120
-90
-60
-30
FiO2 (%)
100
50
0
0
10
Hospital Day
RR-AML_11
Blast Count (%)
100
50
0
-60
-30
FiO2 (%)
100
50
0
0
10
20
30
Hospital Day
Chemotherapy
Hydroxyurea
Prophylactic Antibiotics
Broad Spectrum Antibiotics
Prophylactic Antifungal
Antifungal
Febrile
IPPV
NIPPV
HFNC/Aerosol mask
Nasal Cannula
Bronchoscopy
