## Supplementary figures and images for "Indolent Presentations of Leukemic Lung Disease in Acute Myeloid Leukemia"

### Supplemental Figure E1

Figure E1: Flow diagram of patient selection process

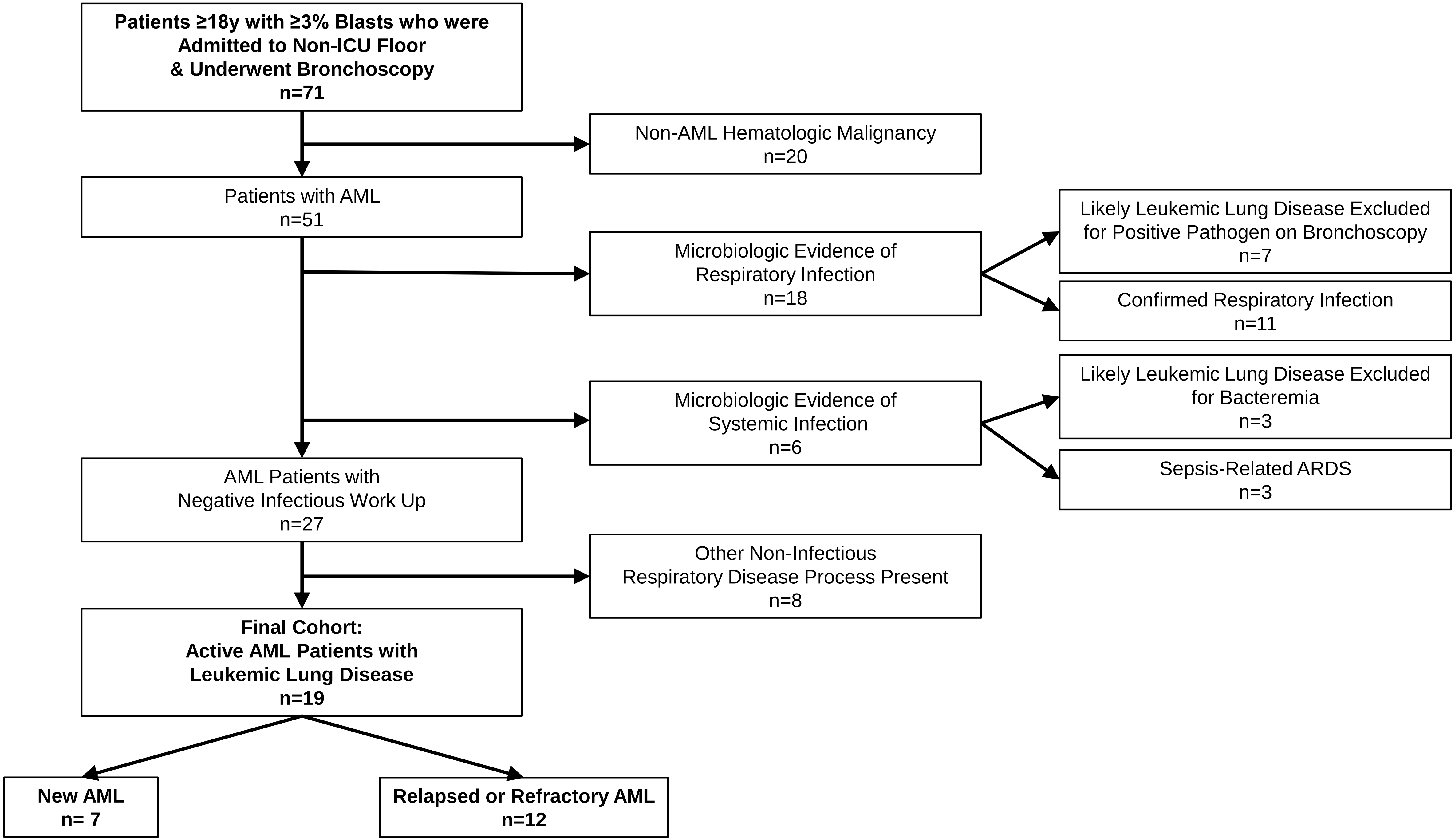
